# Comparative efficacy and acceptability of treatment strategies for antipsychotic-induced akathisia: a systematic review and network meta-analysis

**DOI:** 10.1101/2024.03.06.24303827

**Authors:** Yuki Furukawa, Kota Imai, Yusuke Takahashi, Orestis Efthimiou, Stefan Leucht

## Abstract

**Background:** Antipsychotics are the treatment of choice for schizophrenia, but they often induce akathisia. However, comparative efficacy of treatment strategies for akathisia remains unclear.

**Design:** We performed a systematic review and network meta-analyses (PROSPERO CRD42023450720). We searched multiple databases on 24th July 2023. We included randomized clinical trials comparing one or more treatment strategies for antipsychotic-induced akathisia against each other or control conditions. We included adults with schizophrenia or other psychiatric disorders treated with antipsychotics. The primary outcome was akathisia severity at posttreatment. Secondary outcomes included akathisia response, all-cause dropout, psychotic symptoms, and long-term akathisia severity. We synthesized data in random effects frequentist network meta-analyses and assessed confidence in the evidence using CINeMA.

**Results:** We identified 19 trials with 661 randomized participants (mean age 35.9 [standard deviation 12.0]; 36.7 % [195 of 532] women). No trials examined dose-reduction or switching of antipsychotics. Findings suggested 5-HT2A antagonists (k=6, n=108; standardized mean difference [SMD] -1.07 [95% confidence interval, -1.42; -0.71]) and beta-blockers (k=8, n=105; SMD -0.46 [-0.85; -0.07]) may improve akathisia severity, but confidence in the evidence was deemed low. We also found that benzodiazepines (k=2, n=13; SMD -1.62 [-2.64; -0.59]) and vitamin B6 (k=3, n=67; SMD -0.99 [-1.49; -0.50]) might also be beneficial, but confidence in the evidence was very low. Analyses of secondary outcomes did not provide additional insights.

**Conclusions:** Our findings suggest that 5-HT2A antagonists, beta blockers, and with a lesser certainty, benzodiazepines and vitamin B6 might improve akathisia. These conclusions are extremely preliminary and further trials are needed.

## INTRODUCTION

Antipsychotics are key drugs for treating schizophrenia, but almost one in five patients experience akathisia as a side effect.^1,2^ Akathisia is very irritating, sometimes urges patients to commit harmful behaviors including suicide.^1,3^ Clinical practice guidelines recommend dose reduction, switching antipsychotic, and adjuvant medications.^4,5^ Some adjuvant medications have been examined in a few randomized trials and conventional pairwise meta-analyses.^6^ While the exact pathophysiological mechanism of akathisia remains unclear, the use of these add-on agents is based on hypotheses that antipsychotic-induced akathisia is caused by dopaminergic or serotonergic mechanisms. Anti-parkinsonian drugs, such as anticholinergics and antihistamines are used because akathisia is categorized as an extrapyramidal symptom and suspected to be related to dopaminergic neurotransmission. Beta-blockers may also exert their effects via the inhibitory effect of noradrenergic input to the dopamine system.^7^ 5-HT2A antagonists and triptans may improve akathisia via serotonergic neurotransmission.^8,9^ Benzodiazepines and vitamin B6 are expected to be beneficial because they are shown effective in other movement disorders.^10,11^ To the best of our knowledge, no network meta-analysis of empirical evidence has been performed and the comparative efficacy and acceptability of available treatment strategies remain unknown. In this study, we examined the comparative efficacy and acceptability of treatment strategies for antipsychotic-induced akathisia.

## METHODS

We followed the Preferred Reporting Items for Systematic reviews and Meta-Analyses (PRISMA) guideline extension for NMA.^12^ The protocol was prospectively registered in PROSPERO (CRD42023450720) and can be found in eAppendix1.

### Data sources

#### Criteria for considering studies for this review

We included randomized controlled trials comparing the treatment strategies for antipsychotic-induced akathisia against control conditions. In cross-over trials, we included only the first intervention period to avoid carry-over effects. We included trials on patients using antipsychotics of both sexes aged 18 years or older with schizophrenia or other psychiatric disorders.^13^ We included all drug-related interventions, such as dose reduction of the antipsychotic, switching the antipsychotic, and adjunctive medications such as 5-HT2A antagonists, anticholinergics, antihistamines, benzodiazepines and beta-blockers. (Table 1) We excluded drugs in development and included only licensed drugs.

**TABLE 1.** Classification of the interventions.

| Interventions | Drugs |
| --- | --- |
| **Antipsychotic adjustments** |  |
| Dose reduction | Any antipsychotics |
| Switching | Any antipsychotics |

| Add-on agents |  |
| --- | --- |
| 5-HT2A antagonists | Mianserin, mirtazapine, trazodone |
| Anticholinergics | Benztropine, biperiden, trihexyphenidyl |
| Antihistamines | Cyproheptadine, diphenhydramine, promethazine |
| Benzodiazepines | Clonazepam, diazepam, lorazepam |
| Beta-blockers | Betaxolol, metoprolol, nadolol, propranolol |
| Triptans | Zolmitriptan |
| Vitamin B | Vitamin B6 |

#### Search methods for identification of studies

We searched ClinicalTrials.gov, Cochrane Library (Cochrane Database of Systematic Reviews and Cochrane Central Register of Controlled Trials), Embase, MEDLINE and PsycINFO via Ovid SP, PubMed, and WHO ICTRP on 24th July 2023 with no date, language, document type, and publication status restrictions. Search strategies were developed by a medical information scientist and are reported in eAppendix2. We checked the reference lists of review articles for additional potentially eligible records.

#### Data collection and analysis Selection of studies

Pairs of two reviewers (YF, KI, YT) independently screened titles and abstracts of all the potential studies. We retrieved full text study reports of potentially eligible studies and pairs of two reviewers (YF, KI, YT) independently screened them. We resolved any disagreement through discussion. We identified publications from the same trial so that each trial rather than each report was the unit of analysis in the review. We assessed the inter-rater reliability of the full text screening decisions with Cohen’s κ and percentage agreement.

#### Data items

Pairs of two reviewers (YF, KI, YT) extracted data from the included studies independently. We assessed risk of bias of the primary outcome of the included trials using the revised risk of bias tool by Cochrane^14^ in the five following domains: randomization process, deviations from intended interventions, missing outcome data, measurement of the outcome, and selection of reported results. Any disagreement was resolved through discussion. We measured the inter-rater reliability of the extracted data concerning the primary outcome with intra-class correlation, and the risk of bias assessment with weighted κ and percentage agreement.

#### Primary outcome and secondary outcomes

The primary outcome of interest in this study was treatment efficacy at treatment endpoint. As we prespecified in the protocol, we prioritized the Barnes Akathisia Scale global scale and then total score.^15^ If not available, other validated scales were used. Secondary outcomes included efficacy using akathisia severity response (dichotomous), all-cause dropouts (as a proxy measure of treatment acceptability), psychotic symptom severity and efficacy at long-term follow-up (continuous, longest follow-up between 1 to 12 months). Intention-to-treat analysis was prioritized whenever available. We used standardized mean difference for continuous outcomes, because multiple scales were used, and odds ratio for dichotomous outcomes.^16^

#### Statistical analysis

We created a network diagram at the class-level to visualize the available evidence. Classification of the interventions are described in the Table 1. Transitivity is a basic assumption behind NMA.^17^ To meaningfully combine the direct evidence from A vs C and B vs C studies to learn indirectly about the comparison A vs B, there should not be important differences in the distribution of the effect modifiers across treatment comparisons. We created box plots of trial and patient characteristics deemed to be possible effect modifiers (publication year, proportion of patients with antipsychotics likely to bring akathisia, baseline severity) and visually examined whether they were similarly distributed across treatment comparisons. We checked consistency using global (design-by-treatment) and local (back-calculation) tests.^18,19^ We assessed possible reporting bias and small-study effects using contour-enhanced funnel plots of comparisons with ten or more trials. Given the expected clinical and methodological heterogeneity of treatment effects among the studies, we conducted random-effects NMA. We visualized NMA results using placebo augmentation as reference and ordering treatments considering both the number of participants analyzed and p-scores for the primary outcome, which provide an overall ranking of treatments.^20^ We summarized the results in a table using a coloring scheme where colors denote beneficial or harmful effect and shading shows the strength of confidence in the evidence, which we assessed using the CINeMA approach.^21–23^ We also performed the pairwise created a league table showing NMA results together with direct evidence of pairwise meta-analyses.^23^Finally, we performed a series of pre-specified sensitivity analyses on the primary outcome: 1) we performed individual drug-level NMA to examine the influence of lumping different drugs into classes; 2) we excluded trials with extremely different assessment time point to test the influence of including different endpoint; and 3) we excluded trials with high overall risk of bias to test the influence of risk of bias.^24^

We performed analyses in *R* version 4.2.3^24^ using *netmeta*^25^ and *meta*^26^ packages.

## RESULTS

We identified 6,290 records, assessed 127 full texts for eligibility and included 19 trials with 661 randomized participants in the meta-analysis. (eAppendix2) The inter-rater reliability of judgements for full text screening was good, with κ of 0.67 (95%CI, 0.53-0.81) and percentage agreement of 85%. Appendix lists the excluded trials with reasons for exclusion. (eAppendix2)

Typical participants were males in their thirties (mean age 35.9 [SD 12.0]; 36.7 % [195 of 532] were females). The included trials were small (mean number of participants randomized = 35 [SD 27]). The mean follow-up time was 6.0 (SD 4.3) days. No trial had long-term follow-up longer than one month. No trial examined dose reduction or switching the antipsychotics. Publication year ranged from 1988 to 2020. Majority of the trials were conducted in the Middle East (11 out of 19). Table 2 tabulates the characteristics of included trials. The overall risk of bias according to the Cochrane’s revised risk of bias tool was low in none of the trials, some concerns in seven (37%, 7 out of 19) and high in twelve (63%, 12 out of 19). The inter-rater reliability for the overall risk of bias was slight, with a weighted κ=0.15 (95%CI, 0.00-0.65) and percentage agreement 55%. Inter-rater reliability of extracted primary outcomes was almost perfect, with an intraclass correlation (ICC) of 0.94 (95%CI, 0.88 to 0.98)

**Table 2.**
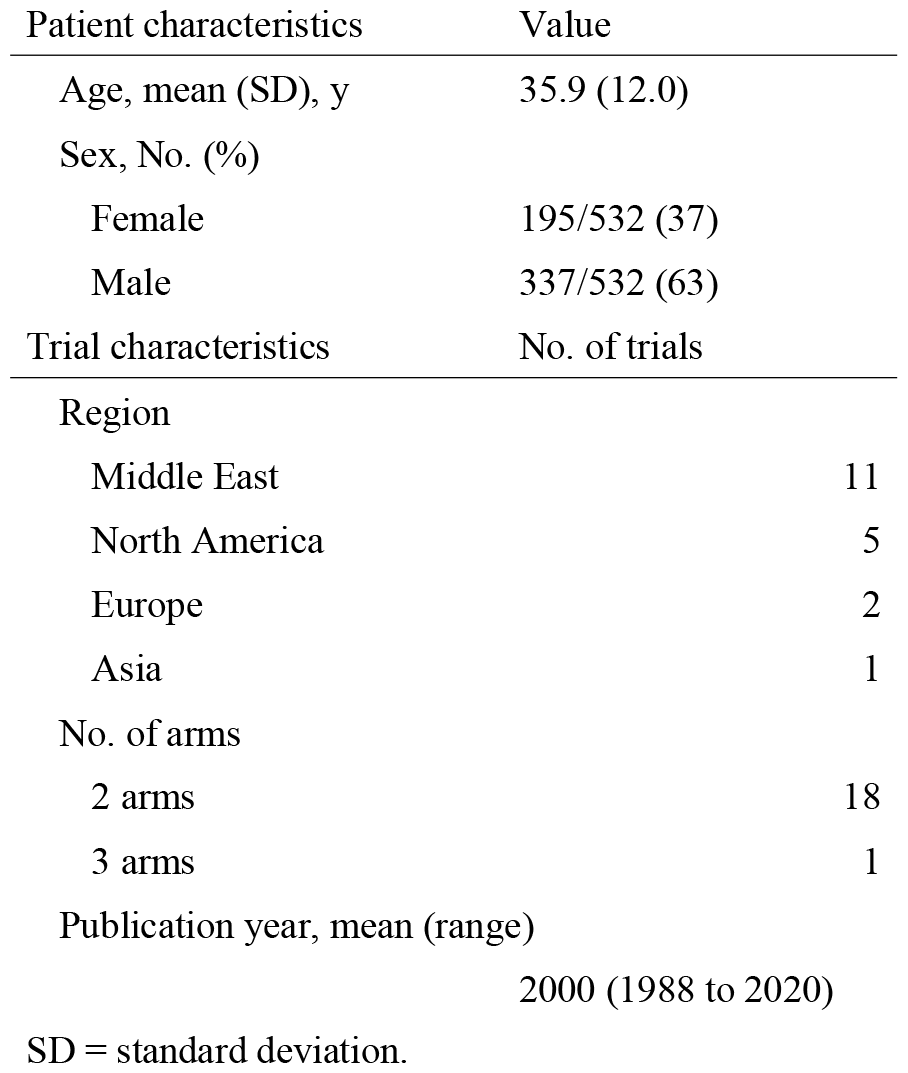
Characteristics of included trials.

| Patient characteristics | Value |
| --- | --- |
| Age, mean (SD), y | 35.9 (12.0) |
| Sex, No. (%) |  |
| Female | 195/532 (37) |
| Male | 337/532 (63) |
| Trial characteristics | No. of trials |
| Region |  |
| Middle East | 11 |
| North America | 5 |
| Europe | 2 |
| Asia | 1 |
| No. of arms |  |
| 2 arms | 18 |
| 3 arms | 1 |
| Publication year, mean (range) |  |
|  | 2000 (1988 to 2020) |

SD = standard deviation.

The network for the primary outcome at the class level was well-connected. (Figure 1) Assessment of transitivity (eAppendix3) found that potential effect modifiers were evenly distributed across comparisons, except for the publication year of beta-blockers and benzodiazepine, which were mainly examined earlier in the 1990s. The global (design-by-treatment) test did not show evidence of inconsistency (p = 0.66). The local (back-calculation) method did not find evidence of disagreement between direct and indirect comparisons (assessed in six comparisons). The limited number of trials precluded an evaluation of publication bias and small-study effects using funnel plots. Figure 2 and Table 3 show the results of the class-level NMA and eAppendix3 shows the results of the pairwise meta-analyses and the league table. eAppendix3 shows the result of CINeMA.^23^ We found that 5-HT2 antagonists (k=6, n=108; standardized mean difference [SMD] -1.07 [95% confidence interval, -1.42 to -0.71]) and beta blockers (k=8, n=105; SMD -0.46 [-0.85 to -0.07]) may be beneficial, but the confidence in the evidence was low according to CINeMA, mainly due to the high overall risk of bias in the original trials. We also found that benzodiazepines (k=2, n=13; SMD -1.62 [-2.64 to -0.59]) and vitamin B6 (k=3, n=67; SMD -0.99 [-1.49 to-0.50]) may be superior to placebo in improving akathisia; confidence in the evidence was, however, very low. Results of the secondary (binary) efficacy outcome were in line with the primary analysis (results in Table3). Dropout for any reason had very wide confidence intervals and did not provide any evidence on the comparative risk of add-on agents (results in Table3). Pre-specified sensitivity analyses were in line with the primary analysis (eAppendix3). We estimated the weighted average proportion of responders in placebo arms to be 16%. Applying the estimated odds ratios of the binary response NMA to this proportion, we estimated that 5-HT2A antagonists may lead to response in 64% (95%CI, 47% to 78%) and beta blockers in 39% (95%CI, 26% to 53%) of the patients.

**Table 3.** Results from network meta-analysis, all outcomes.

|  | Primary outcome |  | Secondary outcomes |  |
| --- | --- | --- | --- | --- |
|  | Akathisia severity | Akathisia response | Dropout | Psychotic symptoms |
| Trials, No. | 17 | 18 | 10 | 10 |
| Heterogeneity |  |  |  |  |
| tau^2 | 0.07 | 0.00 | 0.00 | 0.04 |
| I^2 (95%CI) | 28.5% (0.0%; 63.0%) | 0.0% (0.0%; 55.0%) | 0.0% (0.0%; 79.2%) | 22% (0.0%; 65.1%) |
| Treatments |  |  |  |  |
| Effect measures | SMD (95%CI) | OR (95%CI) | OR (95%CI) | SMD (95%CI) |
| More than 100 participants included for the primary outcome |  |  |  |  |
| 5-HT2A antagonists | -1.07 (-1.42; -0.71) | 9.58 (4.85; 18.9) | 0.56 (0.23; 1.35) | 0.09 (-0.28; 0.45) |
| Beta-blockers | -0.46 (-0.85; -0.07) | 3.38 (1.92; 5.95) | 0.99 (0.39; 2.49) | -0.23 (-0.77; 0.31) |
| Fewer than 100 participants included for the primary outcome |  |  |  |  |
| Benzodiazepines | -1.62 (-2.64; -0.59) | 41.2 (3.97; 428) | 3.00 (0.37; 11.81) | - |
| Vitamin B6 | -0.99 (-1.49; -0.50) | 7.20 (3.06; 17.0) | 0.99 (0.17; 5.78) | -0.21 (-0.75; 0.34) |
| Antihistamines | -0.58 (-1.56; 0.39) | 3.79 (0.78; 18.4) | 2.12 (0.07; 64.2) | 1.10 (0.04; 2.16) |
| Anticholinergics | -0.47 (-1.35; 0.42) | 1.56 (0.36; 6.69) | 3.00 (0.11; 79.5) | -0.39 (-1.22; 0.45) |
| Triptans | 0.03 (-1.05; 1.12) | 1.58 (0.24; 10.5) | - | -0.52 (-1.63; 0.58) |

CI = confidence interval; OR = odds ratio; SMD = standardized mean difference. Light grey indicates treatments being more effective than placebo for beneficial outcomes (akathisia severity and akathisia response), and not more harmful for harmful outcomes (dropout and psychotic symptoms) based on low to very low certainty of evidence according to CINeMA. Dark grey indicates treatments being not better than placebo for beneficial outcomes and more harmful for harmful outcomes based on low to very low certainty of evidence according to CINeMA.

**Figure 1.**
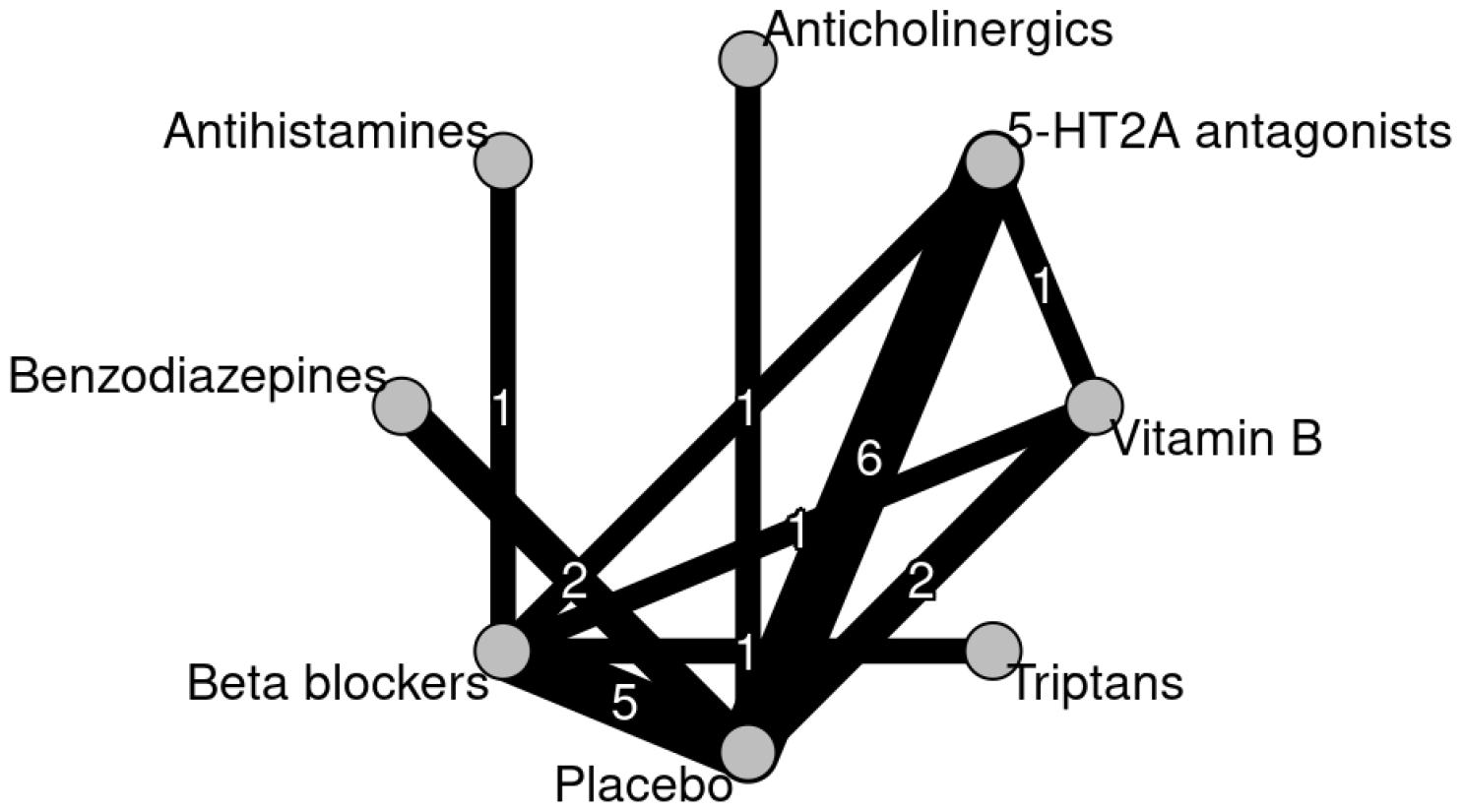
Network diagram.

**Figure 2.**
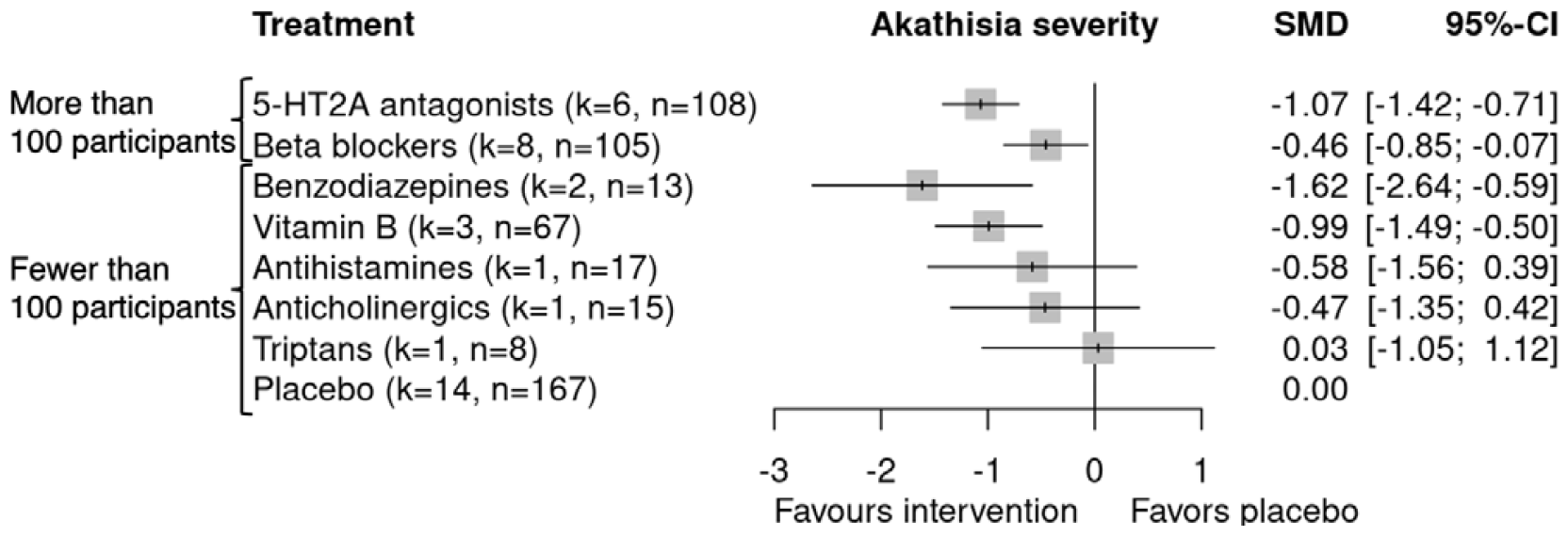
Results from network meta-analysis, for the primary outcome. CI = confidence interval; k denotes the number of arms; n denotes the number of participants analyzed; SMD = standardized mean difference.

## DISCUSSION

To our knowledge, this is the first systematic review and network meta-analysis to examine treatment strategies for antipsychotic-induced akathisia. We found no trials on dose reduction or switching of antipsychotics. There found some evidence suggesting that 5-HT2A antagonists and beta blockers may improve akathisia severity, but the confidence in the evidence was low, owing to the high risk of bias of the original studies, according to our assessment. We also found evidence suggesting that benzodiazepines and vitamin B6 may be also beneficial, but the confidence was very low. These finding are based on short-term follow-up only and no trials examined whether the effects persist on the long term. Analyses of secondary outcomes did not provide additional insight.

Dose-reduction and switching the antipsychotics are the first-line recommendations in clinical practice guidelines.^4,5^ Although we did not find any trials examining these strategies, these strategies remain reasonable; a dose-response meta-analysis of antipsychotics found that higher doses impose a greater risk of akathisia,^27^ and different antipsychotics carry different risks of akathisia.^13^ Albeit the lack of evidence, these strategies should still be considered, given the low to very low confidence in the evidence of add-on agents and the absence of evidence of their long-term efficacy. Clinical practice guideline recommendations for add-on pharmacotherapies vary from guideline to guideline. Some lists all the drugs based on the positive results of a few randomized controlled trials,^6^ while some discourages the use of concomitant agents based on the very limited body of evidence.^5^ A moderate approach is to make a weak recommendation for add-on agents that have at least a little evidence and that are widely used in clinical practice.^4^ This study is the first to address the issue with the most systematic and established way of evidence synthesis.

The strength of our study is the use of network meta-analysis, which allowed us to include five comparisons examining an active intervention to another. These comparisons were excluded in the conventional pairwise meta-analyses comparing an intervention against placebo. This improved the precision of effect estimates and enabled the evaluation of comparative efficacy.

Our study has several limitations. The most notable limitation is the limited number and size of included trials. Even for the most examined agents, there were less than ten trials, with the total number randomized to the agents being only around 100. Second, many of the trials were old and many methodological advances in clinical trials have been made since then. The overall risks of bias were judged to be high in almost two thirds of the trials, with some concerns in the remaining ones. Third, many trials took place before the second-generation antipsychotics became the mainstream approach to treating schizophrenia, and the results may not be completely applicable to the current clinical practice.^4^

## CONCLUSION

Our systematic review and network meta-analysis found no trials on dose-reduction or switching of antipsychotics. We found a possible short-term benefit of adding 5-HT2A antagonists, beta-blockers (low confidence in the evidence), benzodiazepines and vitamin B6 (very low confidence in the evidence). Adjuvant medications may be considered when it is difficult to reduce the dose of or switch the antipsychotic used, acknowledging the limitations in the body of evidence and with careful consideration of side effects. Albeit the lack of evidence, dose-reduction or switching can still be considered, because the previous meta-analyses showed that higher doses of antipsychotic led to an increased risk of akathisia^27^ and that different antipsychotics convey different risks of akathisia.^13^ All these recommendations are extremely preliminary and further well-designed large-scale trials are needed, which can change the results of this study.

## Data Availability

The study used only openly available data. Data and code will be shared upon reasonable request to the corresponding author.

## Patient and public involvement

There was no patient or public involvement in the development of this manuscript.

## Contributors

Dr Farhad Shokraneh, an Information Scientist at Systematic Review Consultants LTD, designed, revised, tested, and ran the search for this review and wrote the methods.

## Acknowledgements

The views expressed are those of the authors and not necessarily those of affiliated organizations.

## Registration

This protocol is prospectively registered in PROSPERO (CRD42023450720).

This research was prospectively registered (#2023178NIe), Ethical Committee, Faculty of Medicine, The University of Tokyo.

## Data sharing

Data and code used for analyses are available on GitHub.

