## Appendix for "Comparative efficacy and acceptability of treatment strategies for antipsychotic-induced akathisia: a systematic review and network meta-analysis"

**eAppendix1** **Protocol**

We registered the protocol in PROSPERO prospectively (CRD42023450720) on 14 August 2023.

**Protocol as of 1 August 2023**

First draft: 2 June 2023

Last updated: 1 August 2023

**Comparative efficacy and acceptability of treatment strategies for antipsychotic-induced akathisia: a protocol for a systematic review and network meta-analysis**

**REVIEW QUESTION**

What is the comparative efficacy and acceptability of treatment strategies for antipsychotic-induced akathisia?

**BACKGROUND**

Antipsychotics are the key drugs for treating schizophrenia, but they often induce akathisia as a side effect.(Musco et al., 2020) Akathisia is very irritating, sometimes urges patients to commit harmful behaviors including suicide.(Bjarke et al., 2022) Clinical practice guidelines recommend dose reduction, switching antipsychotic, and adjuvant medications.(Keepers et al., 2020) Some adjuvant medications have been examined in a few randomized trials and conventional pairwise meta-analyses.(Pringsheim et al., 2018) To the best of our knowledge, however, no network meta-analysis has been performed and the comparative efficacy and acceptability of the treatment strategies remain unknown.

In this study, we will explore the comparative efficacy and acceptability of treatment strategies for antipsychotic-induced akathisia.

**METHODS**

We will follow the Preferred Reporting Items for Systematic reviews and Meta-Analyses (PRISMA) guideline extension for NMA. (Hutton et al., 2015) The protocol will be prospectively registered in PROSPERO. We will record and report any amendments.

**Data sources**

**Criteria for considering studies for this review**

***Study design***

We will include randomized controlled trials comparing one of the treatment strategies for antipsychotic-induced akathisia against control conditions (treatment as usual, placebo augmentation, waiting list, etc). Cluster randomized trials will be included in meta-analyses in accordance with the Cochrane handbook recommendation as long as the allocation was concealed.(Higgins et al., 2022) In cross-over trials, we will only include the first intervention period to avoid carry-over effects.

Participants

We will include trials on patients of both genders aged 18 years or older with schizophrenia or other psychiatric disorders using antipsychotics.(Huhn et al., 2019) We will include trial regardless of the diagnostic criteria used.

***Interventions and controls***

Interventions will include all drug-related interventions, such as dose reduction of the antipsychotic, switching the antipsychotic, and adjunctive medications such as beta-blockers, benzodiazepines, anticholinergics, antihistamines and 5-HT2A antagonists. (Table 1) We will exclude drugs in development and include only licensed drugs. We will not exclude intramuscular injections at this stage, but we may analyze separately if we deem that participants’ characteristics or measurement time points are too different in studies on these interventions as compared to other trials. Control conditions will include, but not limited to, continuation of the antipsychotic already used, waiting list, and placebo augmentation.

TABLE 1 Classification of the interventions

| Class | Drugs |
| --- | --- |
| Dose reduction | Any antipsychotics |
| Switching | Any antipsychotics |
| Beta-blockers | Propranolol, betaxolol, metoprolol, nadolol |
| Benzodiazepines | Clonazepam, lorazepam, diazepam |
| Anticholinergics | Benztropine, biperiden, trihexyphenidyl |
| Antihistamines | Diphenhydramine, promethazine |
| 5-HT2A antagonists | Mianserin, trazodone, mirtazapine |
| Vitamin B6 | Vitamine B6 |

**Search methods for identification of studies**

We will carry out a comprehensive literature search in Embase, MEDLINE, PsycINFO, PubMed and Cochrane Library (CDSR and CENTRAL). We will search “akathisia” in the title and abstract. We will also search ClinicalTrials.gov and WHO International Clinical Trials Registry Platform. We will impose no date, language or publication status restriction. We will check the reference lists of review articles for additional potentially eligible records.

TABLE2 Search strings

| **Database** | **Search strategy/strings** |
| --- | --- |
| ClinicalTrials.gov | Advanced Search  Condition or Disease: Akathisia  Study type: Interventional Studies (Clinical Trials) |
| Cochrane Library (CDSR and CENTRAL) | #1 [mh "Akathisia, Drug-Induced"] OR (Akathisia* OR Pseudoakathisia* OR Acathisia*):ti,ab |
| Embase via Ovid SP | 1 Randomized controlled trial/ or Controlled clinical study/ or randomization/ or intermethod comparison/ or double blind procedure/ or human experiment/ or (random$ or placebo or (open adj label) or ((double or single or doubly or singly) adj (blind or blinded or blindly)) or parallel group$1 or crossover or cross over or ((assign$ or match or matched or allocation) adj5 (alternate or group$1 or intervention$1 or patient$1 or subject$1 or participant$1)) or assigned or allocated or (controlled adj7 (study or design or trial)) or volunteer or volunteers).ti,ab. or (compare or compared or comparison or trial).ti. or ((evaluated or evaluate or evaluating or assessed or assess) and (compare or compared or comparing or comparison)).ab.  2 (random$ adj sampl$ adj7 ("cross section$" or questionnaire$1 or survey$ or database$1)).ti,ab. not (comparative study/ or controlled study/ or randomi?ed controlled.ti,ab. or randomly assigned.ti,ab.)  3 Cross-sectional study/ not (randomized controlled trial/ or controlled clinical study/ or controlled study/ or (randomi?ed controlled or control group$1).ti,ab.)  4 (((case adj control$) and random$) not randomi?ed controlled).ti,ab.  5 (nonrandom$ not random$).ti,ab.  6 ("Random field$" or (random cluster adj3 sampl$)).ti,ab.  7 (review.ab. and review.pt.) not trial.ti.  8 "we searched".ab. and (review.ti. or review.pt.)  9 ("update review" or (databases adj4 searched)).ab.  10 (rat or rats or mouse or mice or swine or porcine or murine or sheep or lambs or pigs or piglets or rabbit or rabbits or cat or cats or dog or dogs or cattle or bovine or monkey or monkeys or trout or marmoset$1).ti. and animal experiment/  11 Animal experiment/ not (human experiment/ or human/)  12 (Systematic review not (trial or study)).ti.  13 or/2-12  14 1 not 13  15 exp Akathisia/ or Drug-Induced Akathisia/ or (Akathisia* or Pseudoakathisia* or Acathisia*).ti,ab.  16 14 and 15  17 limit 16 to embase |
| MEDLINE via Ovid SP | 1 Akathisia, Drug-Induced/ or (Akathisia* or Pseudoakathisia* or Acathisia*).ti,ab.  2 ((Randomized Controlled Trial or Controlled Clinical Trial).pt. or (Randomi?ed or Placebo or Randomly or Trial or Groups).ab. or Drug Therapy.fs.) not (exp Animals/ not Humans.sh.)  3 1 and 2 |
| PsycINFO via Ovid SP | 1 (random* or factorial* or crossover* or cross-over* or placebo* or (doubl* adj blind*) or (singl* adj blind*) or assign* or allocat* or volunteer* or control*).tw.  2 Akathisia/ or (Akathisia* or Pseudoakathisia* or Acathisia*).ti,ab.  3 1 and 2 |
| PubMed (Excluding MEDLINE) | ("Akathisia, Drug-Induced"[MH] OR Akathisia*[TIAB] OR Pseudoakathisia*[TIAB] OR Acathisia*[TIAB]) AND (Randomized Controlled Trial[PT] OR Controlled Clinical Trial[PT] OR Pragmatic Clinical Trial[PT] OR Randomized[TIAB] OR Randomised[TIAB] OR Placebo[TIAB] OR Randomly[TIAB] OR Trial[TIAB] OR Groups[TIAB]) NOT (Animals[MH] NOT Humans[MH]) NOT MEDLINE[SB] |
| WHO ICTRP | Advanced Search  Akathisia in the Condition  Recruitment status is ALL |

**Data collection and analysis**

**Selection of studies**

Two review authors will independently screen titles and abstracts of all the potential studies we identify as a result of the search and code them as ‘retrieve’ or ‘do not retrieve’. We will retrieve the full text study reports/publications and two review authors will independently screen the full text and identify studies for inclusion and identify and record reasons for exclusion of the ineligible studies. We will resolve any disagreement through discussion or, if required, we will consult a third reviewer. We will identify publications from the same study so that each study rather than each report is the unit of analysis in the review. We will record the selection process in sufficient detail to complete a PRISMA flow diagram.

**Data items**

Two review authors will independently extract data from the included studies. Any disagreement will be resolved through discussion, or discussed with a third person if necessary. We will abstract the following information.

***1. Characteristics of the studies***

Name of the study, year of publication, country, study site (single or multi-center), study design (individually randomized or cluster-randomized), population characteristics (mean age, number of women, primary diagnosis, antipsychotic used), intervention (medication, dosing schedule), outcomes (scale used for the primary outcome)

***2. Risk of bias***

We will use Cochrane Risk of Bias 2.0 tool (RoB2) (Sterne et al., 2019) to assess the risk of bias of the primary outcome. We will report the inter-rater agreement in terms of percentage agreement and kappa.

***3. Data to calculate effect sizes***

We will extract data to calculate effect sizes (the number of patients randomized to each arm, the number of patients assessed, the number of responders, the scale used, the mean, standard deviation and the number assessed for continuous outcomes) When only change from baseline to endpoint is reported for continuous outcomes, we will use it instead of endpoint mean.(Costa et al., 2013)

**Primary outcome and secondary outcomes**

The primary outcome of interest in this study is treatment efficacy at endpoint.

1.	Efficacy: akathisia severity (continuous, endpoint)

Secondary outcomes are as follows:

2.	Efficacy using akathisia severity response (dichotomous, endpoint) as the outcome. We will use the treatment response defined by the authors. When it is unavailable, we will define response as 50% or more reduction on the symptom scales and impute response using mean and standard deviation.　(Furukawa et al., 2005)

3.	Acceptability: dropouts for any reason (dichotomous, endpoint)

4.	Psychotic symptoms (continuous, endpoint)

5.	Efficacy at long-term follow-up (continuous, longest follow-up between 1 to 12 months)

Intention-to-treat analysis will be prioritized whenever available. As it is likely that trials use different scales, we will use standardized mean difference for continuous outcomes. We will use the number of participants randomized as the denominator for dichotomous outcomes. We will use odds ratio for dichotomous outcomes.

**Hierarchy of outcome measures**

For efficacy, we will prioritize the Barnes Akathisia Scale global scale and then total score. If not available, the Extrapyramidal Symptom Rating Scale akathisia item, the Drug-Induced Extrapyramidal Symptoms Scale, or any other validated scales. For psychotic symptoms, we will prioritize Positive and Negative Syndrome Scale, and then Brief Psychiatric Rating Scale and, if not available, any other validated scales

**Statistical analysis**

We aim to perform a class-level network meta-analysis. Classification of the interventions are described in the Table 1. Transitivity is an important underlying assumption of the network meta-analysis model.　(Efthimiou et al., 2016) We will examine transitivity by creating a table of important trial and patient characteristics to see if potential effect modifiers (publication year, proportion of patients with antipsychotics likely to bring akathisia (Huhn et al., 2019), baseline severity) are similarly distributed among treatment comparisons. Moreover, for transitivity to hold, we need to ensure that all patients in the network could in principle have received any of the treatments therein. If we deem transitivity to be a plausible assumption, we will proceed with performing a network meta-analysis. Given the expected clinical and methodological heterogeneity of treatment effects among the studies, we will use the random-effects model, assuming a common heterogeneity parameter across the network.

Lack of transitivity may manifest as inconsistency in the network, in case there are closed loops. We will check for consistency using local and global inconsistency tests. If the prerequisites of network meta-analysis are not met, or in case of large unexplained inconsistency, we will only present direct (i.e. from pairwise meta-analyses) and indirect evidence for each treatment comparison.

We will perform all analyses in R (latest version, R foundation, Vienna, Austria) (R_Core_Team, 2020) using netmeta package (Rücker et al., 2020) to conduct network meta-analysis and meta package (Balduzzi et al., 2019) to assess the reporting bias.

**Certainty of evidence**

We will assess the certainty of evidence in network estimates of the primary outcome using CINeMA.(Nikolakopoulou et al., 2020)

**Small study effects and publication bias**

We will assess the presence of small study effects and publication bias, in the evidence set by examining asymmetry in the contour-enhanced funnel plots of all comparisons with ten or more trials using the primary outcome.

**Subgroup analyses**

If we find enough studies, we will investigate the impact of potential effect modifiers by conducting subgroup analyses. We will examine the following characteristics: acute akathisia (begins within the first two weeks of the antipsychotic treatment) vs. tardive akathisia (begins at least three months after the initiation of the antipsychotic treatment), and antipsychotics likely to induce akathisia vs. those less likely to produce akathisia (we will regard those starting from aripiprazole as akathisia-inducing antipsychotics (Huhn et al., 2019)).

**Sensitivity analyses**

We aim to conduct the following sensitivity analyses for the primary outcome only, if data permits.

1.	Drug-level network meta-analysis to see the efficacy of each drug.

2.	Excluding trials with extremely deviated measurement time point

3.	Excluding trials with high overall risk of bias according to RoB2

**Patient and public involvement**

There was no patient or public involvement in the development of this manuscript.

**Acknowledgements**

The views expressed are those of the authors and not necessarily those of affiliated organizations.

**Support**

No financial support will be used.

**Declarations of interest**

YF has received consultancy fee from Panasonic outside the submitted work. SL has received honoraria as a consultant and/or advisor and/or for lectures from Alkermes, Angelini, Eisai, Gedeon Richter, Janssen, Lundbeck, Lundbeck Institute, Merck Sharp and Dohme, Otsuka, Recordati, Rovi, Sanofi Aventis, TEVA, Medichem, Mitsubishi. The other authors declare no interests.

#### Changes from the protocol

14^th^ February, 2024. We decided not to conduct subgroup analyses because we didn’t find enough trials.

### **eAppendix2 Screening process and results**

#### Search and Selection of Studies

We searched ClinicalTrials.gov, Cochrane Library (Cochrane Database of Systematic Reviews and Cochrane Central Register of Controlled Trials=CENTRAL), Embase, MEDLINE and PsycINFO via Ovid SP, PubMed, and WHO ICTRP on 24^th^ July 2023 with no date/time, language, document type, and publication status limitations (protocol). We followed the Cochrane Handbook for Systematic Review of Interventions (1) and Cochrane’s MECIR (2) for conducting the search, PRISMA-S (3), PRISMA guideline (4) for reporting the search, and PRESS guideline for peer-reviewing the search strategies (5). Keywords were collected through experts’ opinions, literature review, controlled vocabulary (APA Thesaurus, Medical Subject Headings = MeSH, and Excerpta Medica Tree = Emtree), and reviewing the primary search results. Because of poor reporting of outcomes in medical research (6-10), we did not limit the search by adding specific outcomes to retrieve all the outcomes.

We used the validated search filters to retrieve RCTs from Embase (11), MEDLINE (1), and PubMed (1). Since there was no validated search filter for PsycINFO, we used a search strategy adapted from CADTH (12). Search strategies developed by a medical information scientist were reported in the protocol. Search results were de-duplicated in EndNote X9 and sent to two researchers for screening. The process of selection of studies has been shown in the PRISMA flow diagram.

1. Lefebvre C, Glanville J, Briscoe S, Littlewood A, Marshall C, Metzendorf MI, et al. Searching for and selecting studies. In: Higgins JPT, Green S, editors. Cochrane Handbook for Systematic Reviews of Interventions. Version 5.1.0 [updated March 2011] ed: The Cochrane Collaboration; 2019. p. 67-107.

2. Higgins JPT, Lasserson T, Chandler J, Tovey D, Churchill R. Methodological Expectations of Cochrane Intervention Reviews. London: Cochrane; 2016.

10. Mantziari S, Demartines N. Poor outcome reporting in medical research; building practice on spoilt grounds. Annals of Translational Medicine. 2017;5(Suppl 1):S15.

11. Glanville J, Foxlee R, Wisniewski S, Noel-Storr A, Edwards M, Dooley G. Translating the Cochrane EMBASE RCT filter from the Ovid interface to Embase.com: a case study. Health Info Libr J. 2019 Jul 22. doi: 10.1111/hir.12269

12. John W. Scott Health Sciences Library. Randomized Controlled Trials / Controlled Clinical Trials: A Cut and Paste Search Strategy adapted from CADTH for Ovid PsycINFO. Adapted from: “Canadian Agency for Drugs and Technology in Health [Internet]. Strings attached: CADTH’s database search filters - randomized controlled trials / controlled clinical trials — OVID Medline, Embase, PsycINFO. Ottawa: CADTH; 2018.” John W. Scott Health Sciences Library, University of Alberta. Rev. December 9, 2020. Available from: https://docs.google.com/document/d/1g7vXZz2CAAqZpbHoOFxxiCVDdCW6bRsDuvqYZDiMH5s/edit.

**Table 1** Search resources details and number of results

| **Resource** | **Time Coverage** | **Search Interface** | **# of Hits** |
| --- | --- | --- | --- |
| ClinicalTrials.gov | Until Search Date | ClinicalTrials.gov | 8 |
| Cochrane Library   - Cochrane Database of Systematic Reviews=CDSR - Cochrane Central Register of Controlled Trials=CENTRAL | Issue 8 of 12, August 2023 | Cochrane Library | 1143 |
| Embase | 1974 – 2023 Week 28 | Ovid SP | 2330 |
| Ovid MEDLINE(R) ALL | 1946 – July 21, 2023 | Ovid SP | 2098 |
| APA PsycINFO | 1806 – July Week 3, 2023 | Ovid SP | 610 |
| PubMed [Excluding MEDLINE] | 1946 – Search Date | PubMed | 89 |
| WHO ICTRP | Until Search Date | WHO ICTRP | 12 |
| Subtotal | 6290 | | |
| Duplicates | 2048 | | |
| **Total (for Screening)** | **4242** | | |

#### PRISMA flow diagram

#### Reports not retrieved

We could not retrieve the following twelve reports. However, by carefully re-assessing the title and abstract and other related papers, we considered them very unlikely to be eligible.

\# Not randomized controlled trials

- Kim A, Adler L, Angrist B, Rotrosen J. Efficacy of low-dose metoprolol in neuroleptic-induced akathisia. J Clin Psychopharmacol. 1989;9(4):294-296.
- Lerner V, Kaptsan A, Miodownik C, Kotler M. Vitamin B6 in treatment of tardive dyskinesia: a preliminary case series study. Clin Neuropharmacol. 1999;22(4):241-243.
- Takahashi H, Kamata M, Yoshida K, Ishigooka J, Higuchi H. Switching to olanzapine after unsuccessful treatment with risperidone during the first episode of schizophrenia: an open-label trial. J Clin Psychiatry. 2006;67(10):1577-1582. doi:10.4088/jcp.v67n1013
- Wehnert A, Stilwell C, Mack R, Sloth-Nielsen M. Extrapyramidal symptoms and sertindole - analysis of three double-blind, Haloperidol-referenced, Phase III clinical trials. 1997. 10th european college of neuropsychopharmacology congress. Vienna, Austria.
- Burgyone K, Aduri K, Ananth J, Parameswaran S. The use of antiparkinsonian agents in the management of drug-induced extrapyramidal symptoms. Curr Pharm Des. 2004;10(18):2239-2248. doi:10.2174/1381612043384123
- Zen Z X, Jing L R, Liu L, Meng Q L, Zhang Y D. Dexetimide and benzhexolum in the treatment of drug-induced akathisia: a double-blind controlled trial. Chinese journal of pharmacoepidemiology. 1995;4(2):72-73. (allocation unclear, according to <https://www.cochranelibrary.com/cdsr/doi/10.1002/14651858.CD003727.pub3/references>)
- Hirose S, Ashby CR. Immediate effect of intravenous diazepam in neuroleptic-induced acute akathisia: an open-label study. J Clin Psychiatry. 2002;63(6):524-527. doi:10.4088/jcp.v63n0610

\# Wrong intervention

- Datta GP. A controlled double blind trial with Trinicalm-plus in schizophrenia. Indian Medical Gazette. 1978;112:141-145.

\# Not enough information in English

- Montoya Cabrera MA, López Martín G, Baca Rodríguez LC, Porcayo Vergara F, Hernández Zamora A, Juárez Aragón G. Síndromes extrapiramidales de evolución aguda causados por drogas [Acute-onset extrapyramidal syndromes caused by drugs]. Bol Med Hosp Infant Mex. 1981;38(4):607-615.
- Schilkrut R, Duran E, Haverbeck C, Katz I, Vidal P. Verlauf von psychopathologischen und extrapyramidalmotorischen Symptomen unter einer Langzeit-Neuroleptikabehandlung schizophrener Patienten [Course of psychopathologic and extrapyramidal motor symptoms during long-term treatment of schizophrenic patients with psycholeptic drugs (author's transl)]. Arzneimittelforschung. 1978;28(9):1494-1495.
- Sikora J, Kabes J, Pisvejc J. Ovlivnĕní nezádoucích úcinků neuroleptik piracetamen Dvojitĕ slepé placebem kontrolované srovnăní [Management of neuroleptic side-effects with piracetam (author's transl)]. Cesk Psychiatr. 1981;77(2):137-142.
- Wang HL. Zhonghua Shen Jing Jing Shen Ke Za Zhi. [Propranolol in the treatment of neuroleptic-induced akathisia]. 1988;21(5):303-320.

#### Excluded reports with reasons (examples)

\# Cross-over trials without the results of the first intervention period alone

- Adler L, Angrist B, Peselow E, Corwin J, Maslansky R, Rotrosen J. A Controlled Assessment of Propranolol in the Treatment of Neuroleptic-Induced Akathisia. *Bri J Psychiatry.* 1986;149(1):42-45. doi:10.1192/bjp.149.1.42
- Sachdev P, Loneragan C. Intravenous benztropine and propranolol challenges in acute neuroleptic-induced akathisia. *Clin Neuropharmacol.* 1993;16(4):324-331. doi:10.1097/00002826-199308000-00004

\# Wrong participants

- Wynchank D, Berk M. Efficacy of nefazodone in the treatment of neuroleptic induced extrapyramidal side effects: a double-blind randomised parallel group placebo-controlled trial. *Hum Psychopharmacol.* 2003;18(4):271-275. doi:10.1002/hup.476

Participants with extrapyramidal side effects other than akathisia were included.

\# Wrong interventions

- Horiguchi J, Nishimatsu O. Usefulness of antiparkinsonian drugs during neuroleptic treatment and the effect of clonazepam on akathisia and parkinsonism occurred after antiparkinsonian drug withdrawal: a double-blind study. *Jpn J Psychiatry Neurol*. 1992;46(3):733-739. doi:10.1111/j.1440-1819.1992.tb00549.x

Control condition (antiparkinsonian drugs) included various drugs and could not be included in this analysis.

\# Ongoing

- IRCT20191218045795N7

\# Unknown status

- NCT00533455
- NCT00659919
- NCT03790345

### **eAppendix3. Network meta-analysis**

#### Assessment of transitivity

###### Publication year

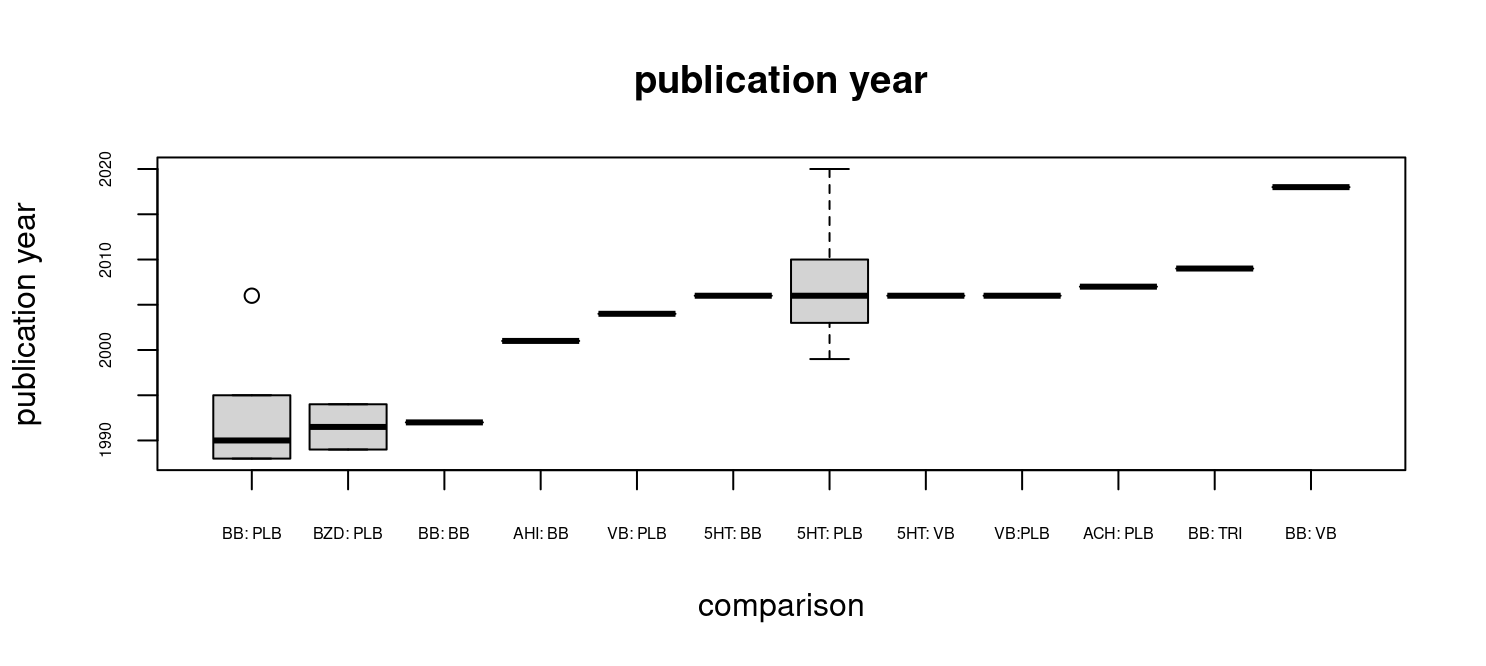

###### Proportion of patients with antipsychotics likely to bring akathisia

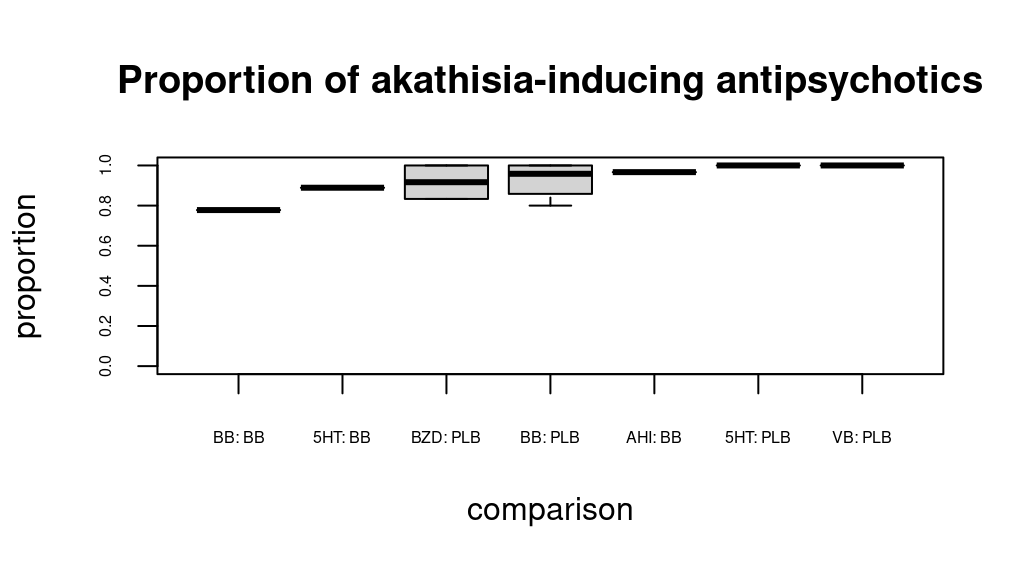

###### Baseline severity (BARS global)

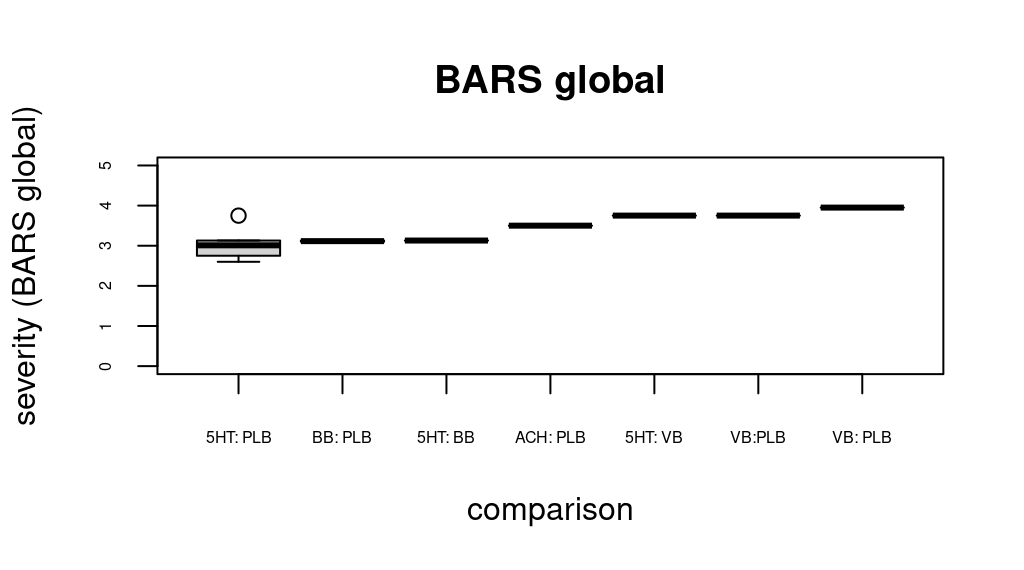

##### global (design-by-treatment)

```
> # global
> decomp.design(net_nma_class)
Q statistics to assess homogeneity / consistency

                    Q df p-value
Total           16.79 12  0.1576
Within designs  10.38  7  0.1680
Between designs  6.41  5  0.2682

Design-specific decomposition of within-designs Q statistic

                     Design    Q df p-value
    placebo:benzodiazepines 8.23  1  0.0041
 placebo:5-HT2A antagonists 1.44  3  0.6953
      placebo:beta blockers 0.70  3  0.8722

Between-designs Q statistic after detaching of single designs
(influential designs have p-value markedly different from 0.2682)

                          Detached design    Q df p-value
 placebo:5-HT2A antagonists:beta blockers 3.16  3  0.3677
     placebo:5-HT2A antagonists:vitamin B 3.31  3  0.3458
                    placebo:beta blockers 4.81  4  0.3078
                  beta blockers:vitamin B 5.65  4  0.2265
               placebo:5-HT2A antagonists 5.84  4  0.2118
                        placebo:vitamin B 6.28  4  0.1789

Q statistic to assess consistency under the assumption of
a full design-by-treatment interaction random effects model

                   Q df p-value tau.within tau2.within
Between designs 3.28  5  0.6567     0.3438      0.1182
```

##### local (back-calculation) test

```
> # local
> netsplit(net_nma_class) # here you can check which (and how many) loops show inconsistencies between direct and indirect TEs.
Separate indirect from direct evidence (SIDE) using back-calculation method

Random effects model:

                          comparison k prop     nma  direct  indir.    Diff     z p-value
 5-HT2A antagonists:anticholinergics 0    0 -0.6023       . -0.6023       .     .       .
    5-HT2A antagonists:antihistamines 0    0 -0.4828       . -0.4828       .     .       .
  5-HT2A antagonists:benzodiazepines 0    0  0.5480       .  0.5480       .     .       .
    5-HT2A antagonists:beta blockers 1 0.42 -0.6097 -0.2728 -0.8565  0.5837  1.22  0.2225
          5-HT2A antagonists:placebo 6 0.93 -1.0676 -1.1125 -0.5049 -0.6076 -0.88  0.3792
         5-HT2A antagonists:triptans 0    0 -1.1002       . -1.1002       .     .       .
        5-HT2A antagonists:vitamin B 1 0.47 -0.0755 -0.1766  0.0129 -0.1895 -0.35  0.7284
      anticholinergics:antihistamines 0    0  0.1195       .  0.1195       .     .       .
    anticholinergics:benzodiazepines 0    0  1.1502       .  1.1502       .     .       .
      anticholinergics:beta blockers 0    0 -0.0075       . -0.0075       .     .       .
            anticholinergics:placebo 1 1.00 -0.4654 -0.4654       .       .     .       .
           anticholinergics:triptans 0    0 -0.4979       . -0.4979       .     .       .
          anticholinergics:vitamin B 0    0  0.5267       .  0.5267       .     .       .
       antihistamines:benzodiazepines 0    0  1.0307       .  1.0307       .     .       .
         antihistamines:beta blockers 1 1.00 -0.1270 -0.1270       .       .     .       .
               antihistamines:placebo 0    0 -0.5849       . -0.5849       .     .       .
              antihistamines:triptans 0    0 -0.6174       . -0.6174       .     .       .
             antihistamines:vitamin B 0    0  0.4072       .  0.4072       .     .       .
       benzodiazepines:beta blockers 0    0 -1.1577       . -1.1577       .     .       .
             benzodiazepines:placebo 2 1.00 -1.6156 -1.6156       .       .     .       .
            benzodiazepines:triptans 0    0 -1.6482       . -1.6482       .     .       .
           benzodiazepines:vitamin B 0    0 -0.6235       . -0.6235       .     .       .
               beta blockers:placebo 5 0.79 -0.4579 -0.2875 -1.0913  0.8038  1.65  0.0996
              beta blockers:triptans 1 1.00 -0.4905 -0.4905       .       .     .       .
             beta blockers:vitamin B 1 0.46  0.5342  0.3034  0.7319 -0.4285 -0.80  0.4231
                    triptans:placebo 0    0  0.0325       .  0.0325       .     .       .
                   vitamin B:placebo 2 0.56 -0.9921 -1.3043 -0.5866 -0.7177 -1.40  0.1602
                  triptans:vitamin B 0    0  1.0246       .  1.0246       .     .       .

Legend:
 comparison - Treatment comparison
 k          - Number of studies providing direct evidence
 prop       - Direct evidence proportion
 nma        - Estimated treatment effect (SMD) in network meta-analysis
 direct     - Estimated treatment effect (SMD) derived from direct evidence
 indir.     - Estimated treatment effect (SMD) derived from indirect evidence
 Diff       - Difference between direct and indirect treatment estimates
 z          - z-value of test for disagreement (direct versus indirect)
 p-value    - p-value of test for disagreement (direct versus indirect)
```

#### CINeMA

We considered reporting bias of all the comparisons to be “some concerns.” We regarded SMD 0.5 as clinically important effect size. We downgraded the comparisons with number of studies less than 3. We downgraded one level with one domain of “major concerns” and one, two and three domains of “some concerns.”

| **Comparison** | **Number of studies** | **Within-study bias** | **Reporting bias** | **Indirectness** | **Imprecision** | **Heterogeneity** | **Incoherence** | **Confidence rating** |
| --- | --- | --- | --- | --- | --- | --- | --- | --- |
| **5-HT2A antagonists:beta blockers** | 1 | Major concerns | Some concerns | -- | No concerns | Some concerns | No concerns | Very low |
| **5-HT2A antagonists:placebo** | 6 | Major concerns | Some concerns | -- | No concerns | No concerns | No concerns | Low |
| **5-HT2A antagonists:vitamin B** | 1 | Some concerns | Some concerns | -- | Some concerns | Some concerns | No concerns | Very low |
| **anticholinergics:placebo** | 1 | Some concerns | Some concerns | -- | Some concerns | Some concerns | No concerns | Very low |
| **antihistamines:beta blockers** | 1 | Major concerns | Some concerns | -- | Major concerns | No concerns | No concerns | Very low |
| **benzodiazepines:placebo** | 2 | Major concerns | Some concerns | -- | No concerns | No concerns | No concerns | Very low |
| **beta blockers:placebo** | 5 | Major concerns | Some concerns | -- | No concerns | Some concerns | No concerns | Low |
| **beta blockers:triptans** | 1 | Major concerns | Some concerns | -- | Some concerns | Some concerns | No concerns | Very low |
| **beta blockers:vitamin B** | 1 | Major concerns | Some concerns | -- | No concerns | Some concerns | No concerns | Very low |
| **placebo:vitamin B** | 2 | Some concerns | Some concerns | -- | No concerns | No concerns | No concerns | Very low |
| **5-HT2A antagonists:anticholinergics** | 0 | Some concerns | Some concerns | -- | Some concerns | No concerns | No concerns | Very low |
| **5-HT2A antagonists:antihistamines** | 0 | Major concerns | Some concerns | -- | Some concerns | Some concerns | No concerns | Very low |
| **5-HT2A antagonists:benzodiazepines** | 0 | Major concerns | Some concerns | -- | Major concerns | No concerns | No concerns | Very low |
| **5-HT2A antagonists:triptans** | 0 | Major concerns | Some concerns | -- | No concerns | Some concerns | No concerns | Very low |
| **anticholinergics:antihistamines** | 0 | Major concerns | Some concerns | -- | Major concerns | No concerns | No concerns | Very low |
| **anticholinergics:benzodiazepines** | 0 | Some concerns | Some concerns | -- | Some concerns | Some concerns | No concerns | Very low |
| **anticholinergics:beta blockers** | 0 | Some concerns | Some concerns | -- | Major concerns | No concerns | No concerns | Very low |
| **anticholinergics:triptans** | 0 | Major concerns | Some concerns | -- | Major concerns | No concerns | No concerns | Very low |
| **anticholinergics:vitamin B** | 0 | Some concerns | Some concerns | -- | Some concerns | Some concerns | No concerns | Very low |
| **antihistamines:benzodiazepines** | 0 | Major concerns | Some concerns | -- | Major concerns | No concerns | No concerns | Very low |
| **antihistamines:placebo** | 0 | Major concerns | Some concerns | -- | Some concerns | No concerns | No concerns | Very low |
| **antihistamines:triptans** | 0 | Major concerns | Some concerns | -- | Major concerns | No concerns | No concerns | Very low |
| **antihistamines:vitamin B** | 0 | Major concerns | Some concerns | -- | Major concerns | No concerns | No concerns | Very low |
| **benzodiazepines:beta blockers** | 0 | Major concerns | Some concerns | -- | Some concerns | No concerns | No concerns | Very low |
| **benzodiazepines:triptans** | 0 | Major concerns | Some concerns | -- | No concerns | Some concerns | No concerns | Very low |
| **benzodiazepines:vitamin B** | 0 | Major concerns | Some concerns | -- | Major concerns | No concerns | No concerns | Very low |
| **placebo:triptans** | 0 | Major concerns | Some concerns | -- | Major concerns | No concerns | No concerns | Very low |
| **triptans:vitamin B** | 0 | Major concerns | Some concerns | -- | Some concerns | No concerns | No concerns | Very low |

#### Forest plots for secondary outcomes

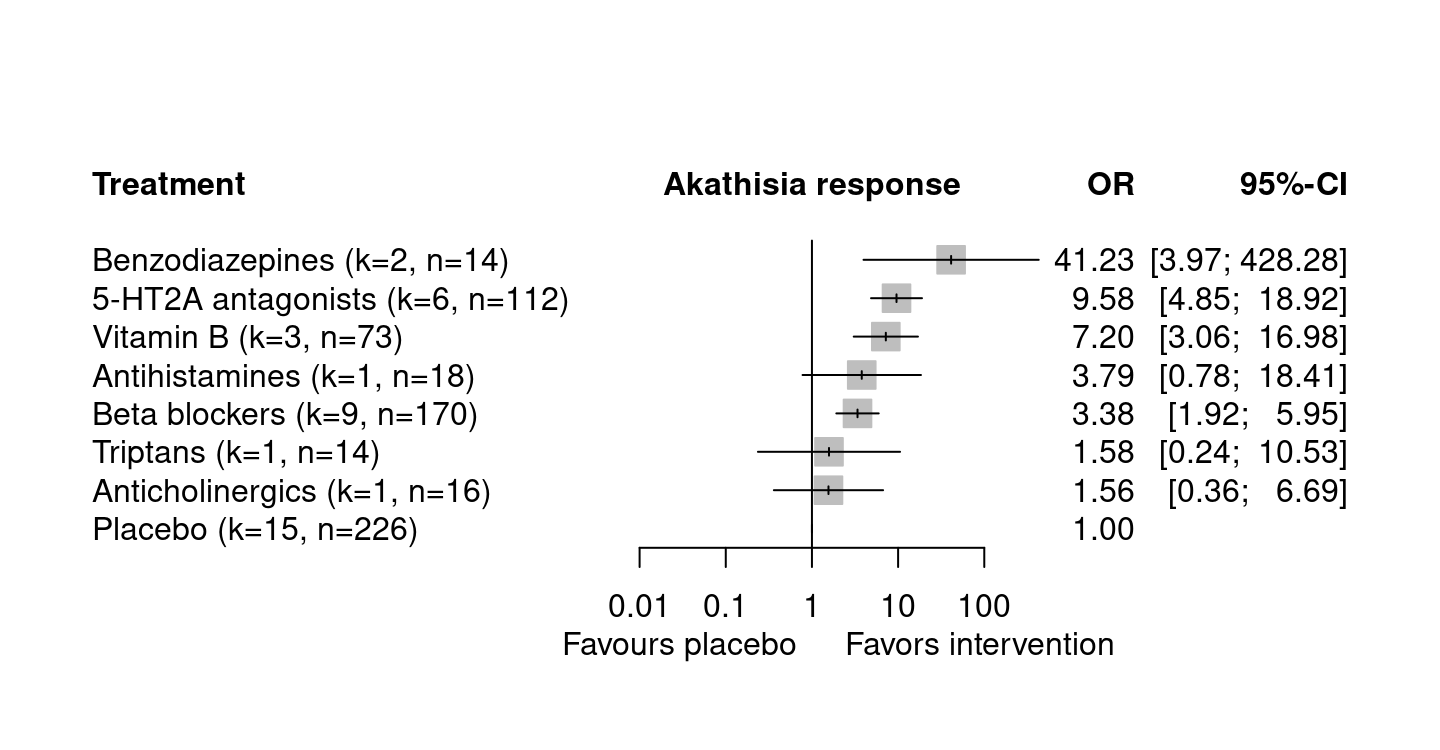

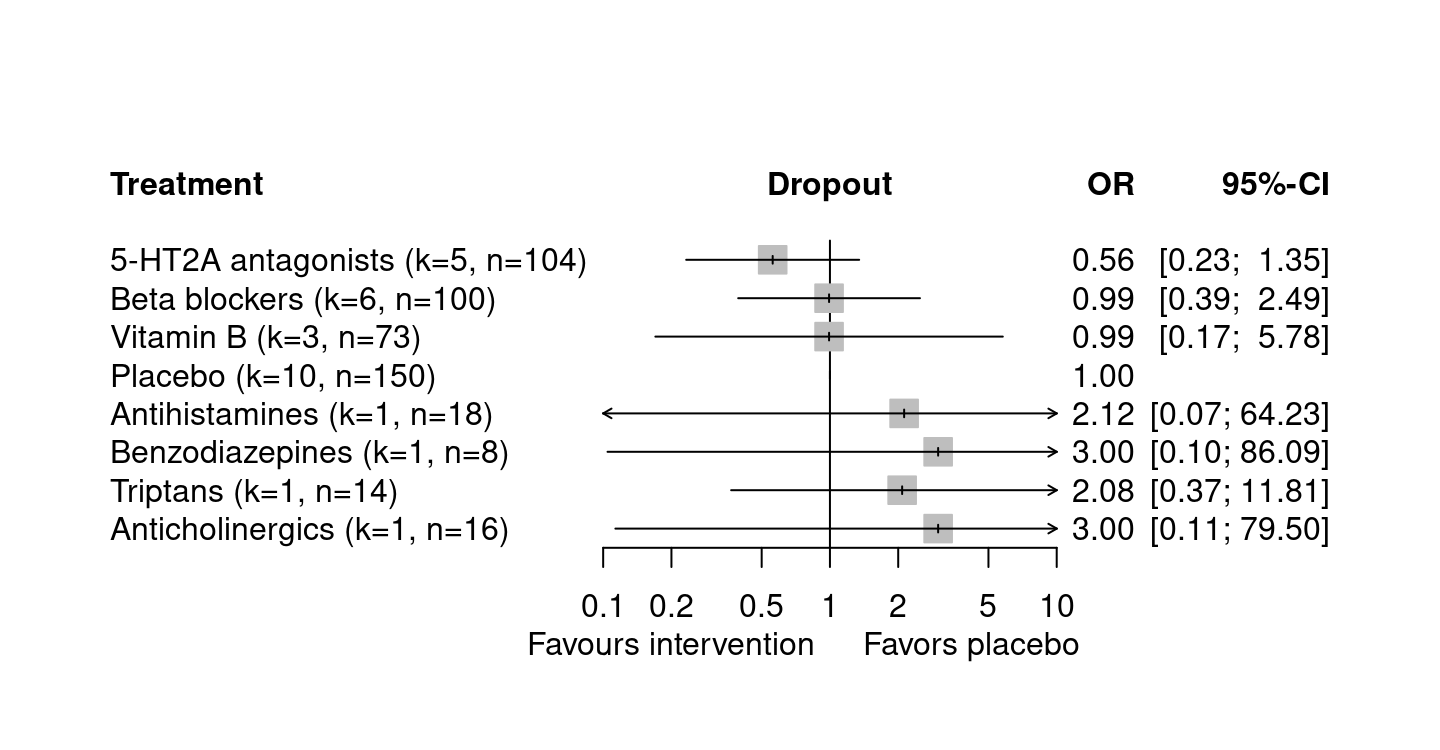

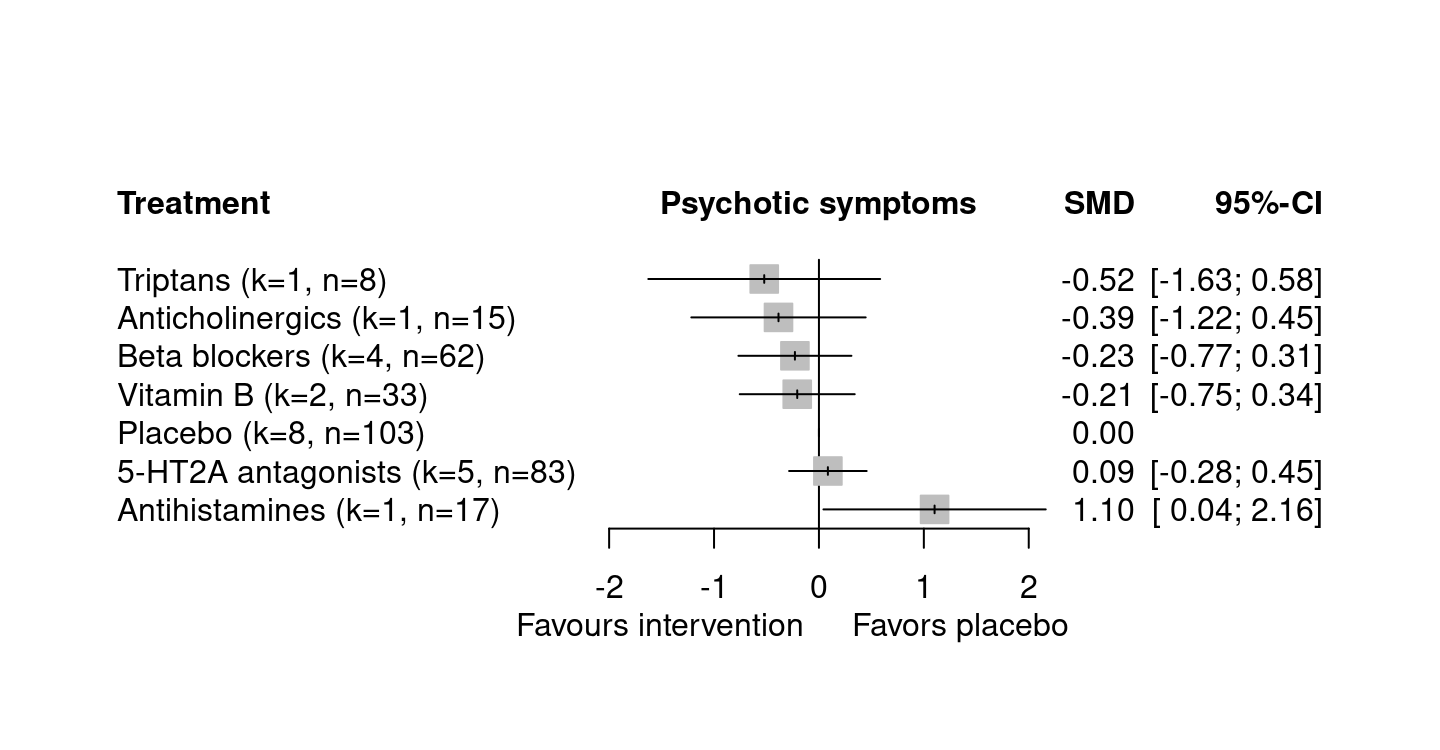

K denotes the number of arms; n denotes the number of participants analyzed.

#### Sensitivity analyses

We aim to conduct the following sensitivity analyses for the primary outcome only, if data permits.

1. Drug-level network meta-analysis to see the efficacy of each drug.

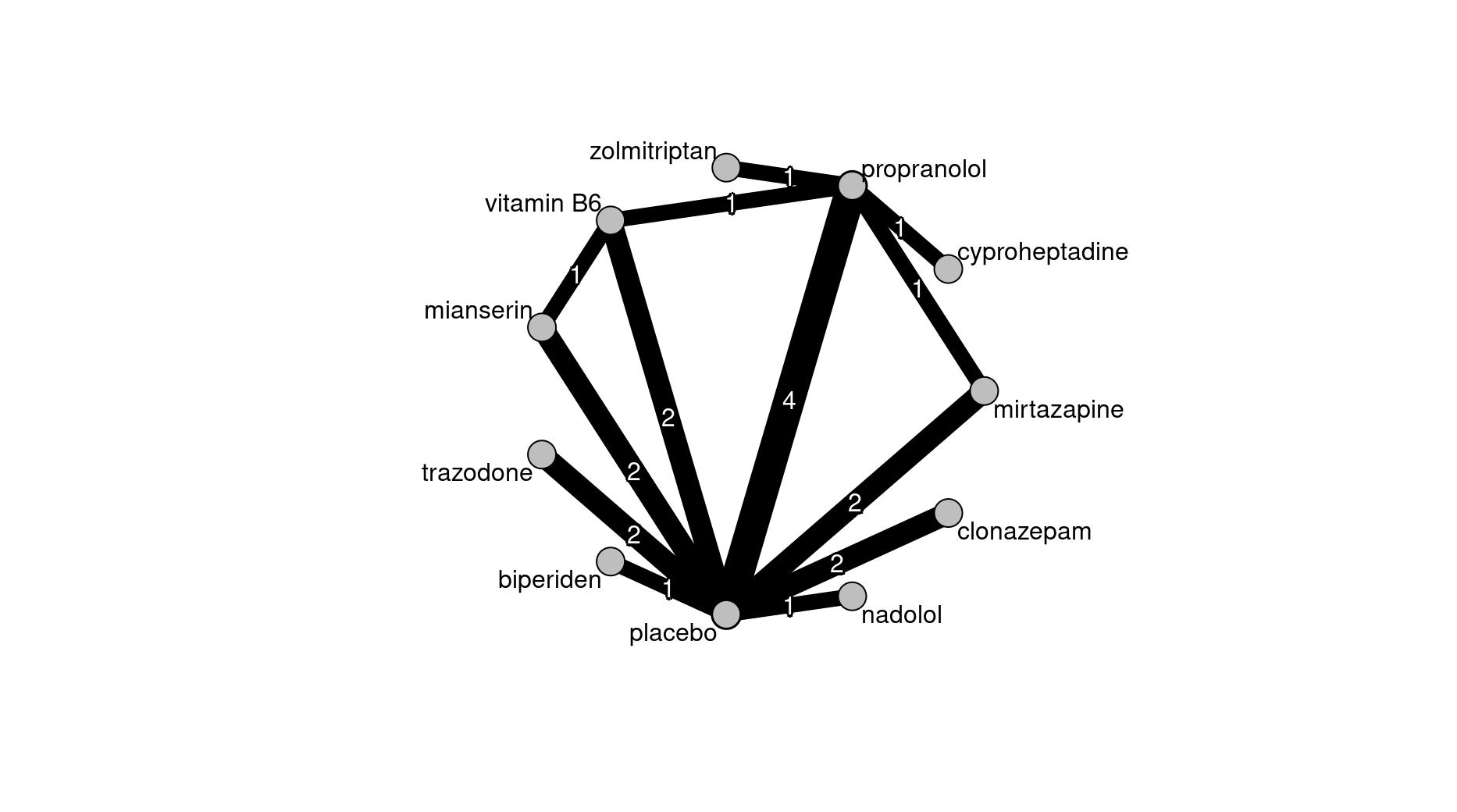

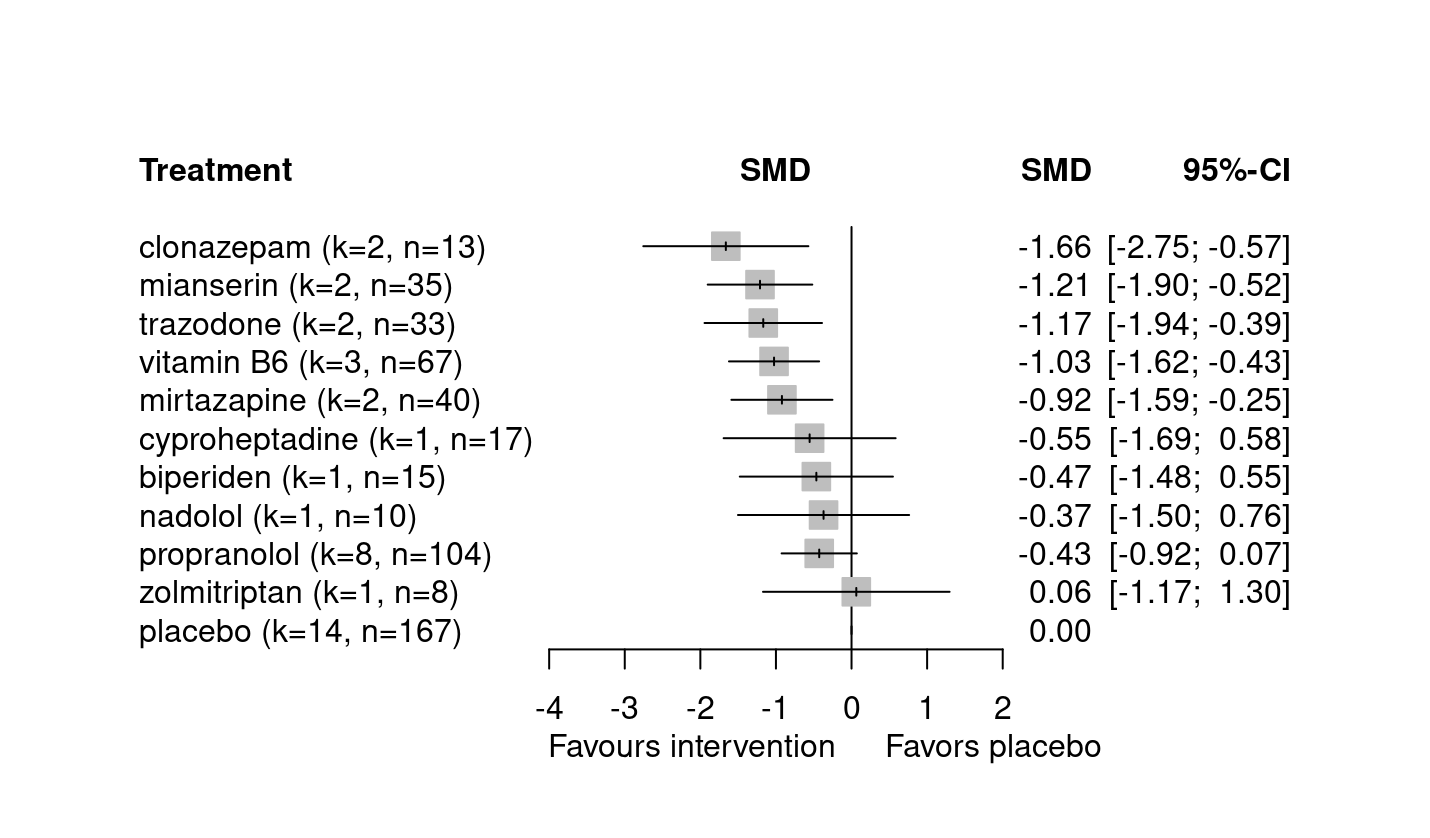

2. Excluding trials with extremely deviated measurement time point

We excluded Baskak2007, where they measured the primary outcome in less than a day.

3.	Excluding trials with high overall risk of bias according to RoB2

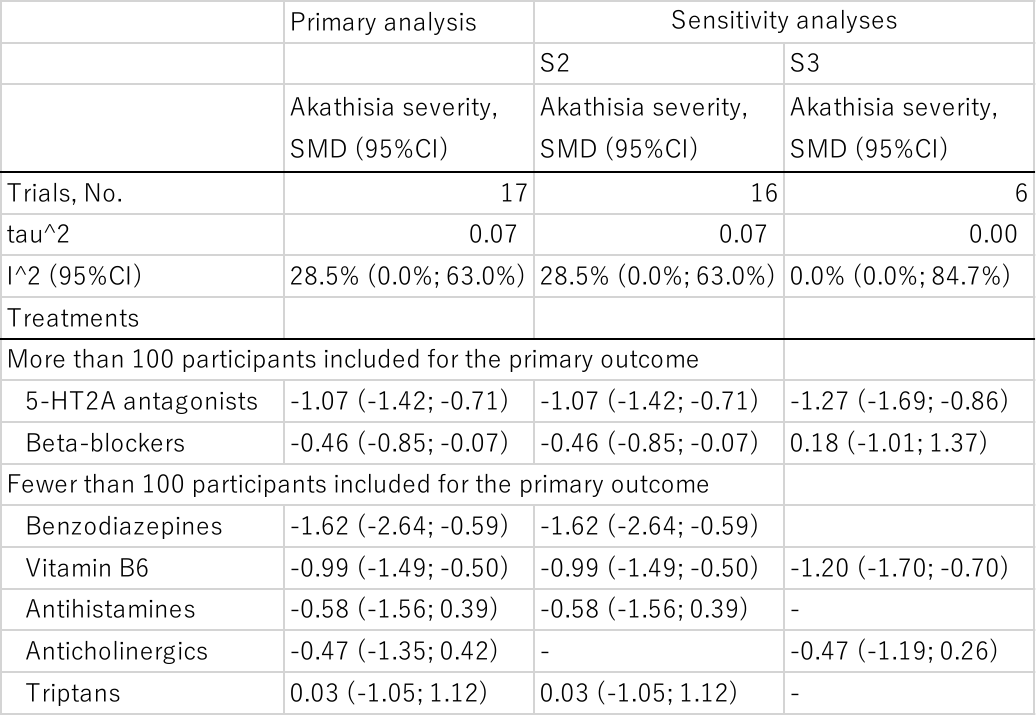

#### Pairwise meta-analyses

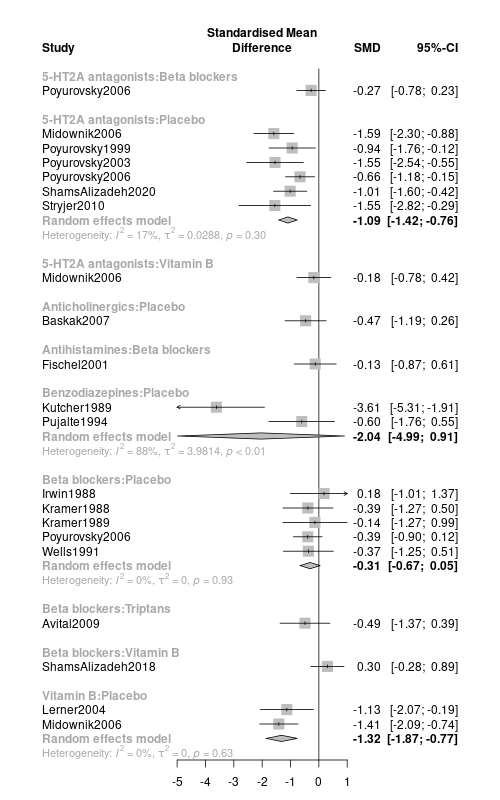

#### League table

| Benzodiazepines | . | . | . | . | . | . | -1.62  (-2.64 to -0.59) (k=2, n=26) |
| --- | --- | --- | --- | --- | --- | --- | --- |
| -0.55  (-1.63 to 0.54) | 5-HT2A antagonists | -0.18  (-0.96 to 0.60) (k=1, n=43) | . | . | -0.27 (-0.99 to 0.44) (k=1, n=60) | . | -1.11  (-1.48 to -0.74) (k=6, n=206) |
| -0.62  (-1.76 to 0.52) | -0.08  (-0.61 to 0.46) | Vitamin B | . | . | -0.30  (-1.07 to 0.47) (k=1, n=51) | . | -1.30  (-1.96 to -0.64) (k=2, n=60) |
| -1.03  (-2.45 to 0.39) | -0.48  (-1.49 to 0.52) | -0.41  (-1.44 to 0.63) | Antihistamines | . | -0.13  (-1.02 to 0.77) (k=1, n=29) | . | . |
| -1.15  (-2.50 to 0.20) | -0.60  (-1.55 to 0.35) | -0.53  (-1.54 to 0.48) | -0.12  (-1.43 to 1.19) | Anticholinergics | . | . | -0.47  (-1.35 to 0.42) (k=1, n=30) |
| -1.16  (-2.26 to -0.06) | -0.61  (-1.07 to -0.15) | -0.53  (-1.06 to -0.01) | -0.13  (-1.02 to 0.77) | -0.01  (-0.97 to 0.96) | Beta blockers | -0.49  (-1.50 to 0.52) (k=1, n=51) | -0.29  (-0.73 to 0.15) (k=5, n=123) |
| -1.65  (-3.14 to -0.15) | -1.10  (-2.21 to 0.01) | -1.02  (-2.16 to 0.11) | -0.62  (-1.97 to 0.73) | -0.50  (-1.90 to 0.90) | -0.49  (-1.50 to 0.52) | Triptans | . |
| -1.62  (-2.64 to -0.59) | -1.07  (-1.42 to -0.71) | -0.99  (-1.49 to -0.50) | -0.58  (-1.56 to 0.39) | -0.47  (-1.35 to 0.42) | -0.46  (-0.85 to -0.07) | 0.03  (-1.05 to 1.12) | Placebo |

Network meta-analysis results in the bottom-left corner. Direct evidence in the upper-right corner. K denotes the number of comparisons; n denotes the number of participants analyzed in the comparison.

### **eAppendix4 PRISAMA-NMA**

| **Section/Topic** | **Item #** | **Checklist Item** | **Reported on Page #** |
| --- | --- | --- | --- |
| **TITLE** |  |  |  |
| Title | 1 | Identify the report as a systematic review *incorporating a network meta-analysis (or related form of meta-analysis).* | P1 |
| **ABSTRACT** |  |  |  |
| Structured summary | 2 | Provide a structured summary including, as applicable:  **Background:** main objectives  **Methods:** data sources; study eligibility criteria, participants, and interventions; study appraisal; and *synthesis methods, such as network meta-analysis.*  **Results:** number of studies and participants identified; summary estimates with corresponding confidence/credible intervals; *treatment rankings may also be discussed. Authors may choose to summarize pairwise comparisons against a chosen treatment included in their analyses for brevity.*  **Discussion/Conclusions:** limitations; conclusions and implications of findings.  **Other:** primary source of funding; systematic review registration number with registry name. | P2 |
| **INTRODUCTION** |  |  |  |
| Rationale | 3 | Describe the rationale for the review in the context of what is already known*, including mention of why a network meta-analysis has been conducted.* | P3 |
| Objectives | 4 | Provide an explicit statement of questions being addressed, with reference to participants, interventions, comparisons, outcomes, and study design (PICOS). | P3 |
| **METHODS** |  |  |  |
| Protocol and registration | 5 | Indicate whether a review protocol exists and if and where it can be accessed (e.g., Web address); and, if available, provide registration information, including registration number. | P3, appendix |
| Eligibility criteria | 6 | Specify study characteristics (e.g., PICOS, length of follow-up) and report characteristics (e.g., years considered, language, publication status) used as criteria for eligibility, giving rationale. *Clearly describe eligible treatments included in the treatment network, and note whether any have been clustered or merged into the same node (with justification).* | P3 |
| Information sources | 7 | Describe all information sources (e.g., databases with dates of coverage, contact with study authors to identify additional studies) in the search and date last searched. | P4 |
| Search | 8 | Present full electronic search strategy for at least one database, including any limits used, such that it could be repeated. | Appendix |
| Study selection | 9 | State the process for selecting studies (i.e., screening, eligibility, included in systematic review, and, if applicable, included in the meta-analysis). | P4, Appendix |
| Data collection process | 10 | Describe method of data extraction from reports (e.g., piloted forms, independently, in duplicate) and any processes for obtaining and confirming data from investigators. | P4 |
| Data items | 11 | List and define all variables for which data were sought (e.g., PICOS, funding sources) and any assumptions and simplifications made. | Appendix |
| **Geometry of the network** | **S1** | Describe methods used to explore the geometry of the treatment network under study and potential biases related to it. This should include how the evidence base has been graphically summarized for presentation, and what characteristics were compiled and used to describe the evidence base to readers. | Figure1 |
| Risk of bias within individual studies | 12 | Describe methods used for assessing risk of bias of individual studies (including specification of whether this was done at the study or outcome level), and how this information is to be used in any data synthesis. | Table 2 |
| Summary measures | 13 | State the principal summary measures (e.g., risk ratio, difference in means). *Also describe the use of additional summary measures assessed, such as treatment rankings and surface under the cumulative ranking curve (SUCRA) values, as well as modified approaches used to present summary findings from meta-analyses.* | P4-5 |
| Planned methods of analysis | 14 | Describe the methods of handling data and combining results of studies for each network meta-analysis. This should include, but not be limited to:   - *Handling of multi-arm trials;* - *Selection of variance structure;* - *Selection of prior distributions in Bayesian analyses; and* - *Assessment of model fit.* | P4-5 |
| **Assessment of Inconsistency** | **S2** | Describe the statistical methods used to evaluate the agreement of direct and indirect evidence in the treatment network(s) studied. Describe efforts taken to address its presence when found. | P4-5 |
| Risk of bias across studies | 15 | Specify any assessment of risk of bias that may affect the cumulative evidence (e.g., publication bias, selective reporting within studies). | P4-5 |
| Additional analyses | 16 | Describe methods of additional analyses if done, indicating which were pre-specified. This may include, but not be limited to, the following:   - Sensitivity or subgroup analyses; - Meta-regression analyses; - *Alternative formulations of the treatment network; and* - *Use of alternative prior distributions for Bayesian analyses (if applicable).* | P5 |
| **RESULTS†** |  |  |  |
| Study selection | 17 | Give numbers of studies screened, assessed for eligibility, and included in the review, with reasons for exclusions at each stage, ideally with a flow diagram. | Appendix |
| **Presentation of network structure** | **S3** | Provide a network graph of the included studies to enable visualization of the geometry of the treatment network. | Appendix |
| **Summary of network geometry** | **S4** | Provide a brief overview of characteristics of the treatment network. This may include commentary on the abundance of trials and randomized patients for the different interventions and pairwise comparisons in the network, gaps of evidence in the treatment network, and potential biases reflected by the network structure. | P5 |
| Study characteristics | 18 | For each study, present characteristics for which data were extracted (e.g., study size, PICOS, follow-up period) and provide the citations. | Table 2 |
| Risk of bias within studies | 19 | Present data on risk of bias of each study and, if available, any outcome level assessment. | Table 2 |
| Results of individual studies | 20 | For all outcomes considered (benefits or harms), present, for each study: 1) simple summary data for each intervention group, and 2) effect estimates and confidence intervals. *Modified approaches may be needed to deal with information from larger networks.* | Appendix |
| Synthesis of results | 21 | Present results of each meta-analysis done, including confidence/credible intervals. *In larger networks, authors may focus on comparisons versus a particular comparator (e.g. placebo or standard care), with full findings presented in an appendix. League tables and forest plots may be considered to summarize pairwise comparisons.* If additional summary measures were explored (such as treatment rankings), these should also be presented. | Figure2, Table3, Appendix |
| **Exploration for inconsistency** | **S5** | Describe results from investigations of inconsistency. This may include such information as measures of model fit to compare consistency and inconsistency models, *P* values from statistical tests, or summary of inconsistency estimates from different parts of the treatment network. | Appendix |
| Risk of bias across studies | 22 | Present results of any assessment of risk of bias across studies for the evidence base being studied. | Appendix CINeMA |
| Results of additional analyses | 23 | Give results of additional analyses, if done (e.g., sensitivity or subgroup analyses, meta-regression analyses*, alternative network geometries studied, alternative choice of prior distributions for Bayesian analyses,* and so forth). | Appendix |
| **DISCUSSION** |  |  |  |
| Summary of evidence | 24 | Summarize the main findings, including the strength of evidence for each main outcome; consider their relevance to key groups (e.g., healthcare providers, users, and policy-makers). | P10 |
| Limitations | 25 | Discuss limitations at study and outcome level (e.g., risk of bias), and at review level (e.g., incomplete retrieval of identified research, reporting bias). *Comment on the validity of the assumptions, such as transitivity and consistency. Comment on any concerns regarding network geometry (e.g., avoidance of certain comparisons).* | P10 |
| Conclusions | 26 | Provide a general interpretation of the results in the context of other evidence, and implications for future research. | P10 |
| **FUNDING** |  |  |  |
| Funding | 27 | Describe sources of funding for the systematic review and other support (e.g., supply of data); role of funders for the systematic review. This should also include information regarding whether funding has been received from manufacturers of treatments in the network and/or whether some of the authors are content experts with professional conflicts of interest that could affect use of treatments in the network. | P11 |
